# AN OVERVIEW OF THE IMPACT OF SOCIAL SUPPORT ON NURSE BURN OUT IN HOSPITALS

**DOI:** 10.1101/2023.12.10.23299642

**Authors:** Silvia Dewi Mayasari Riu, Moses Glorino Rumambo Pandin

## Abstract

**Introduction:** The nursing profession is one of the most difficult and stressful occupations. Nurses have a great responsibility in providing health care to clients, which often involves emotional, complex, and high-risk situations. Social support is emotional, informational and practical assistance provided by individuals or groups to others in certain situations. Burnout syndrome is a chronic stress condition that affects many individuals including nurses. This condition often arises as a result of excessive workload, high physical and emotional demands and imbalance between work and personal life plus the occurrence of a global pandemic.

**Method:** The author used the literature review method by searching for references via the internet. The literature search used Sciencedirect, Pubmed, and Medline and Google Scholar with keywords in English: “social support”, “burnout”. The inclusion criteria in this research were articles that discussed social support on burnout, were in English, published in the last 5 years (2018-2023), explored in the article was the influence of social support on burnout, and a comprehensive examination of the full article is provided. Exclusion criteria were articles that did not discuss social support on burnout.

**Result:** From the literature search, it was obtained 16 journal articles that were relevant with the research objectives.

**Conclusion:** Social support has an important role as a protective factor that can reduce the risk of burnout in individuals, especially among professionals or workers who experience high workloads. Good social support can provide the emotional, instrumental, and informational resources needed to cope with daily stress and pressure.

**Suggestion:** Social support can be considered a form of “natural protection” against burnout, and efforts to strengthen social support networks can be an effective strategy in preventing and coping burnout at various levels of life and professions.

## INTRODUCTION

The nursing profession is one of the most difficult and stressful occupations. Nurses have a great responsibility in providing health care to clients, which often involves emotional, complex, and high-risk situations. In carrying out their duties, nurses are often faced with situations that require quick decisions, caring for sick or injured clients, interacting with anxious clients’ families, especially during a global pandemic. These pressures can cause nurses to experience stress, burnout, and negative impacts on nurses’ well-being. Social support for nurses is currently needed because the level of work pressure for nurses in the last three years has increased sharply. One of the challenges often faced by nurses is Burnout Syndrome. It is a chronic stress condition that affects many individuals including nurses. This condition often arises as a result of excessive workload, high physical and emotional demands, and imbalance between work and personal life plus the occurance of a global pandemic (Solon et al., 2021). The substantial problem lies in nurse burnout, given its potential to negatively impact the physical and mental health of nurses, ultimately jeopardizing the quality of care extended to patients. (John Wiley, 2017).

Data obtained that nurses who experience burnout in the world are 47.9%, this prevalence shows that burnout in nurses is very high (Nursing CE Central 2021). To overcome burnout, it is important for nurses and health facility managers to take preventive measures and provide each other with the necessary psychological support and resources power required. This includes stress management, peer support, psychological assistance, and reasonable restrictions on working hours. Recovery from burnout can take time, and it’s important to recognize the early signs so that action can be taken as quickly as possible. Burnout in nurses is not only an individual problem, but also has an impact on patients and the performance of nurses provided (Albazoon et al., 2023).

Social support is emotional, informational, or practical assistance provided by individual or group to another person in a certain situation. Social support has an important role in maintaining a person’s psychological and physical well-being, as well as helping individuals cope with stress, pressure or difficulties in their lives. Social support has a very important urgency in the lives of individuals and society in general. Here are some reasons why social support is indispensable: it improves emotional well-being and helps individuals overcome stress, anxiety and depression and help individuals feel happier.

Reduces Stress, Social support helps individuals deal with stress better. People who feel supported are better able to deal with stress and change in their lives (Bry & Wigert, 2022).

Improved physical health, Research has shown that having a strong social support network can contribute to better physical health. Support from social networks can decrease the likelihood of developing heart disease, reduce blood pressure, and enhance the functioning of the immune system. (Ruiz-fern et al., 2021).

Improved adaptability, Social support helps individuals adapt to changes in life, this allows individuals to recover more quickly and find more effective solutions.

Furthermore, increased self-confidence. Support from others, particularly in the form of positive encouragement and acknowledgment, has the potential to enhance an individual’s self-assurance and motivation to accomplish objectives and surmount challenges.

Reduced risk of substance abuse. Strong social support can help prevent substance abuse, as individuals who feel supported are more likely to seek support and understanding rather than relying on illicit substances as an escape.

Improved Social Relationships Social support can help strengthen relationships between individuals, family and friends. This can create stronger bonds within the community and society.

Improved workplace productivity and well-being at work. Social support at work can improve employee productivity, job satisfaction and employee well-being. Employees who feel supported tend to be more productive and contribute positively to the work environment.

Overcoming feelings of loneliness and isolation. Support from others plays a crucial role in alleviating sensations of loneliness and social isolation, both of which can detrimentally affect mental and physical well-being. Social support has a wide-reaching impact on various aspects of an individual’s life, and a lack of social support can lead to mental and physical health problems.

In this context, social support becomes an important factor in helping nurses overcome nurse burnout. Social support can encompass a range of elements, such as emotional assistance, informative aid, affiliative backing, and pragmatic assistance. However, social support can be used as a medium to reduce burnout in nurses.

## METHOD

The literature search for this research used articles in English originating from Sciencedirect, Pubmed, and Medline and Google Scholar from 2018 to 2023. The literature search used the key words “social support”, “burnout nurse”, you can see based on the image Flow diagram below.

Prisma Flow chart diagram

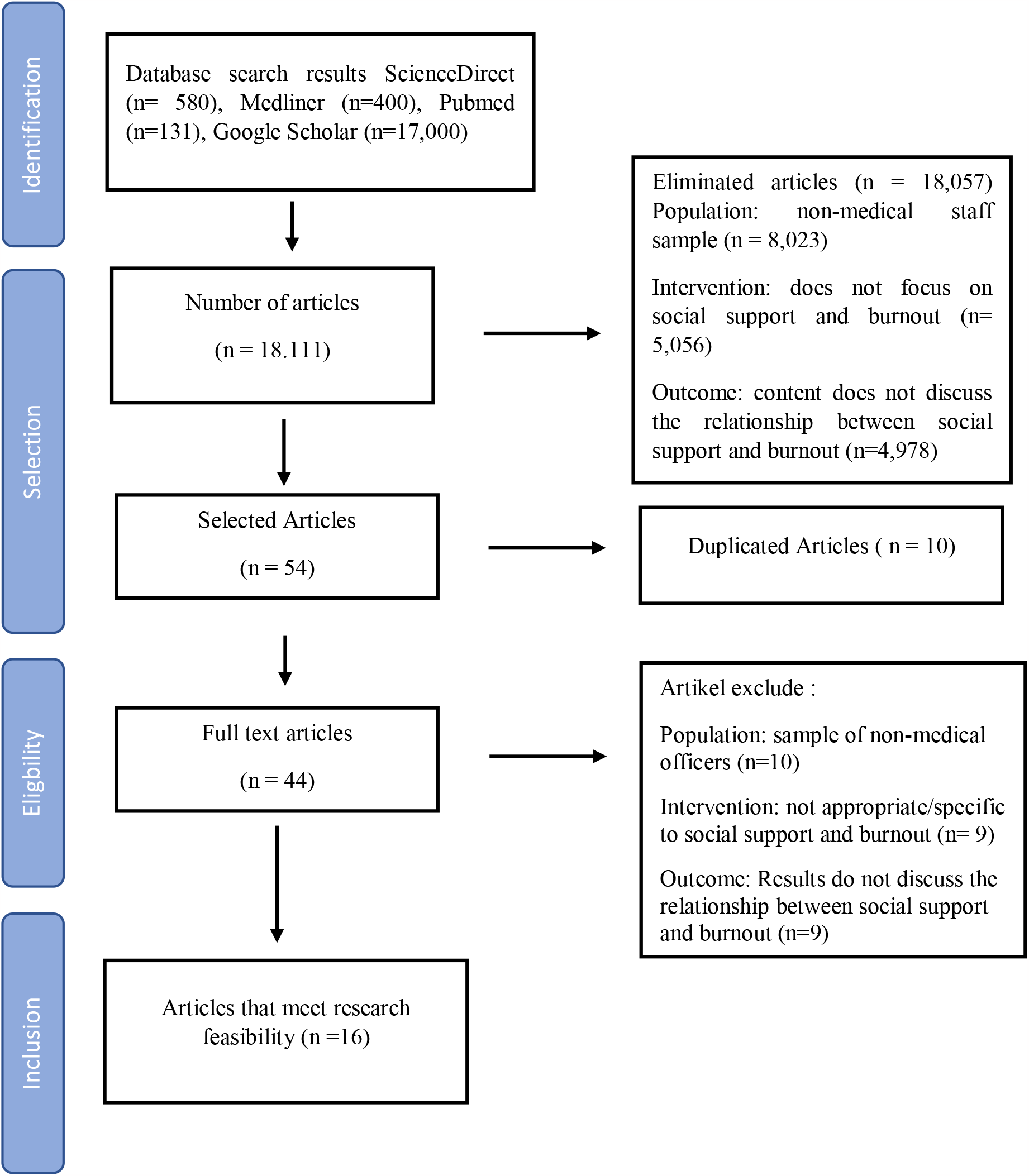

search results : ScienceDirect (n= 580), Medliner (n=400), Pubmed (n=131), Google Scholar (n=17,000). Number of articles obtained (n = 18,111), articles eliminated (n = 18,057)

Population: sample No officer medical (n=8,023) Intervention: no focuses on support social and burnout (n= 5,056) Outcome: content does not discuss related between social support and burnout (n=4,978), Exclude articles: *Population* : sample No officer medical (*n* =10) *Intervention* : no appropriate / specific with support social and burnout (*n* = 9) *Outcome* : Results not discussed related between social support and burnout (*n* =9), Corresponding articles with appropriateness research (n =16).

**Table 1:** List of Journals in the Included Category of the PRISMA Flow.

| No | Title , Author , Year | Method | Results |
| --- | --- | --- | --- |
| 1 | Burnout and post-traumatic stress disorder symptoms among medical staff two years after the COVID-19 pandemic in Wuhan, China: Social support and resilience as mediators<br>Liu et al., 2023 | **Design** : Cross-Sectional<br>**Subject** : Hospital Health Staff<br>**Variables** : Burnout, Social Support, Resilience<br>**Instruments** : MBI-GS, MSPSS, PCL-5<br>**Analysis**: ANOVA | The existence of social support and psychological resilience exhibited a distinct and significant negative relationship with burnout and symptoms associated with post-traumatic stress disorder (PTSD), respectively. (P < 0.001). |
| 2 | Social Support Outside the Workplace, Coping Styles, and Burnout in a Cohort of EMS Providers From Minnesota<br>Boland et al., 2019 | **Design** : Cross-Sectional<br>**Subject** : EMS Provider<br>**Variables**: Burnout, Social support<br>**Instrument**: MBI<br>**Analysis**: Logistic Regression Model | As this research on EMS organizations shows, surveys assessing mental well-being indicators including job burnout, perceived stress, PTSD symptoms, and suicidal tendencies are interrelated. |
| 3 | Emergency room nurses' pathway to turnover intention: a moderated serial mediation analysis<br>(Bruyneel et al., 2016) | **Design** : Cross-sectional<br>**Subjects**: 292 nurses<br>**Variables**: work demands, skills, decision making, social support, quality of work environment, burnout<br>**Instruments** : LQWQ-N, PES-NWI, MBI-HSS<br>**Analysis** : SEM | The findings of the study indicate that robust backing from supervisors and colleagues, augmented decision-making autonomy, and enhanced skill knowledge have a mitigating impact on the occurrence of emotional exhaustion. |
| 4 | The impact of social support and stress on academic burnout among medical students in online learning: The mediating role of resilience<br>Liu & Cao, 2022 | **Design** : Cross-Sectional<br>**Subject** : Medical student<br>**Variables**: Social Support, Burnout, Stress, Resilience<br>**Instruments**: MSPSS, MBI-SS<br>**Analysis**: Linear Regression | The results of this study suggest that academic burnout is predicted to increase with both stress and resilience. However, there is no significant association between social support and academic burnout. |
| 5 | Burnout, Social Support, and Job Satisfaction among Jordanian Mental Health Nurses<br>(Hamaideh, 2011) | **Design** : Cross-Sectional<br>**Subject** : 181 nurses<br>**Variables**: Burnout, Social Support, Job Satisfaction<br>**Instruments**: MBI, SSS, JSS<br>**Analysis**: Multiple Linear Regression | Social support may to some extent have a protective effect on burnout and poor relationships with coworkers result in burnout. |
| 6 | Moderated Role of Social Support in the Relationship between Job Strain, Burnout, and Organizational Commitment among Operating Room Nurses: A Cross-Sectional Study<br>Li et al., 2022 | **Design :** Cross-Sectional<br>**Subject :** 509 nurse<br>**Variables :** Social Support , Job Strain, Burnout, Organizational Commitment<br>**Instruments :** Chinese version of the Job Content Questionnaire, Maslach Burnout Inventory-General Survey<br>**Analysis :** Multiple-group Path | High and low social support groups where social support can moderate the impact of job strain, fatigue and organizational commitment. |
| 7 | The Association between Korean Clinical Nurses' Workplace Bullying, Positive Psychological Capital, and Social Support on Burnout<br>Bae et al., 2021 | **Design:** Cross – Sectional<br>**Subjects :** 166 nurses<br>**Variables :** Social Support, Burnout, Bullying<br>**Instruments :** Korean Version Tedium Scale, WPBN-TI, Korean Version Positive Psychological Capital tool, Korean Version Social Support tool<br>**Analysis :** Hierarchical Regression, T-Tests And ANOVA, Spearman Correlation | The results of the analysis demonstrate a direct association between burnout and bullying, underscoring a link between these elements. Conversely, there is an inverse relationship between burnout and both psychological capital and supportive social environments, suggesting that elevated levels of psychological capital and favorable social settings correlate with reduced levels of burnout. |
| 8 | The mediating role of social support in the relationship between physician burnout and professionalism behaviors (Song et al., 2021) | **Design :** Cross – Sectional<br>**Subject :** 3506 Doctor<br>**Variables :** Professionalism , Burnout, Social Support<br>**Instruments:** BMPI, MBI-2, SSRS<br>**Analysis:** Linear Regression and Logistic Regression | The research findings revealed a negative association between burnout and professionalism, particularly concerning aspects of respect and responsibility. Nevertheless, there was no notable link observed between burnout and either integrity or excellence. Moreover, social support demonstrated a positive correlation with professionalism across all its behavioral dimensions. Additionally, it was identified that social support plays a role as a partial mediator in the relationship between burnout and professionalism. |
| 9 | Occupational and Psychological Factors Associated With Burnout in Night Shift Nurses (Min et al., 2023) | **Design :** Cross-Sectional<br>**Subject :** 231 night shift nurses<br>**Variables:** Burnout, anxiety, patient health, stress, quality of life, social support<br>**Instruments :** MBI-GS, PHQ-9, | study This show that resilience and support more social _ big linked with reduced fatigue emotional , depersonalization , and escalation efficacy |
|  |  | GAD-7, PSS, WHOQOL-BREF, FSSQ<br>**Analysis :** Logistic Regression | professional . |
| 10 | Depression and burnout among Chinese nurses during COVID-19 pandemic: a mediation and moderation analysis model among frontline nurses and nonfrontline nurses caring for COVID-19 patients (Wang et al., 2023) | **Design :** Cross - Sectional<br>**Subject :** 4517 Nurse<br>**Variable:** Burnout, Interpersonal relationships, depression<br>**Instruments:** DASS, MBI-GS<br>**Analysis :** Spearman Colleration | Study This explore possibility mechanism depression caused _ by fatigue , strengthen potency role mediation from style coping positivity and relationships interpersonal ( support social ), as well as give base theoretical For reduce impact Burnout's negative impact on psychological status nurse . |
| 11 | The relationship between psychosocial job stress and burnout in emergency departments: An exploratory study (Mariano et al., 2012) | **Design :** Cross-sectional<br>**Subjects:** 191 emergency room nurses<br>**Variables:** Job Stressors, Burnout<br>**Instruments:** NSS, MBI<br>**Analysis:** PCA | The research results found that interpersonal conflict, lack of social support, employee contract status and excessive workload are the core of burnout. |
| 12 | Social Support Mediates the Effect of Burnout on Health in Health Care Professionals Ruisoto et al., 2021 | **Design :** cross-sectional<br>**Subject :** 608 physicians and 427 nurses<br>**Variables :** social support, burnout,<br>**Instruments :** MOS, MBI<br>**Analysis :** Pearson's tests and t-tests | Study This highlighting that burnout level in energy health can own impact negative on health they in a way overall , but support social can Act as a mitigating mediator impact the . Study results show that exists support strong social _ can help reduce effect bad burnout on health physical and mental professionals health . |
| 13 | Anxiety symptoms and burnout among Chinese medical staff of intensive care units: the moderating effect of social support Zhang et al., 2020 | **Design :** cross-sectional<br>**Subject :** 615 ICU doctors and nurses<br>**Variables :** burnout, social support, symptoms worry<br>**Instruments :** SAS, SSRS, CBI<br>**Analysis :** | Social support can be significant in connecting fatigue to symptoms of anxiety at two distinct stages. |
| 14 | The relationship between social support and burnout among ICU nurses in Shanghai: A cross-sectional study (Li, Ruan & Yuan, 2015) | **Design:** Quantitative<br>**Subjects:** 362 ICU Nurses<br>**Variable:** Social Support<br>**Instrument:** Social Support Questionnaire by Xiao Shui Yuan<br>**Analysis:** Multiple Regression (MRA) | An examination of the social support profiles reveals that nurses receive ample support from their families, but encounter a deficiency in support from colleagues and friends. Nevertheless, they do receive some level of support from their superiors. There is no overlap observed between<br>support from supervisors, family, coworkers, or friends.<br>The study's results indicate a notable prevalence of burnout among intensive care unit (ICU) nurses in Shanghai, China. Our findings propose that factors such as supervisor support, coworker support, taking leave, and work demands significantly contribute to emotional exhaustion. Key factors influencing depersonalization include support from supervisors, age, and job demands. Enhancing personal achievement may be achieved through increased support from supervisors. It is imperative for future research and interventions to address not only individual factors but also extend their impact to broader social and environmental aspects. |
| 15 | Relationships Among Character Strengths, Self-efficacy, Social Support, Depression, and Psychological Well-being of Hospital Nurses ( Xie et al, 2020 ) | **Design :** Quantitative<br>**Subject :** 364 Nurses<br>**Variables :** strength character , support social , depression , and well-being psychological<br>**Instruments :** Social Support Self-Rating Scale (SSRS )<br>**Analysis :** Structural equation modeling (SEM) | There is a relationship between strength character , support social , depression , and well-being psychological . Strength character help people build and maintain connection social , acquiring a sense of belonging social , and improve support social they . Our research show that nurses in China own level support social ones moderate , that is direct and positive influenced by power character they . Nurse with not quite enough answer professional and family Want to No Want to must face work conflict family . Support social important for nurse ; matter This can lighten up<br>pressure them and improve performance work they . Apart from that , nurses with level social support which are more tall Possible experience achievement more personal _ high and fatigue more emotional _ A little . Whereas social support influential direct and negative to depression and positively affects mental health. Hence, the presence of positive character traits eases the burden of depression and enhances mental well-being by fostering social assistance. |
| 16 | Workplace Social Support as a Mediating Factor in the Association between Occupational Stressors and Job Burnout: A Study in the Taiwanese Nursing Context (Wu, Chou & Kao, 2023) | **Design:** Quantitative<br>**Subjects:** 516 Nurses<br>**Variables:** Social Support , work stress<br>**Instruments :** Job Content Questionnaire (C-JCQ)<br>**Analysis:** Structural equation modeling (SEM) | The challenges confronted by nurses and the support they receive in the workplace contribute to job burnout. Social support plays a potential mediating role in the connection between nurses' stressors and burnout. This research reveals a positive correlation between social support, reduced nurse stressors, and decreased job burnout. Enhanced organizational support, positive managerial relationships, and effective patient care contribute to the improvement of nurse practitioner practice. Furthermore, studies indicate that higher levels of organizational support are linked to increased job satisfaction among nurses. In the context of Taiwan's rapid development, nurses grapple with a demanding work environment influenced by an aging population and shifting disease patterns. The rising healthcare demands and patient volumes necessitate nurses to acquire advanced skills and devote more time<br>to their clinical duties. This heightened workload can lead to emotional fatigue and reduced personal gratification. Considering the potential impact of work burnout on patient care quality in healthcare settings, assessing burnout levels among nurses can raise awareness about the challenges faced by these compassionate professionals. |

## Results

Overall, the literature review shows a significant relationship between social support and burnout levels. Social support has an important role as a protective factor that can reduce the risk of burnout in individuals, especially among professionals or workers who experience high workloads. Good social support can provide the emotional, instrumental, and informational resources necessary to cope with everyday stress and pressure. In many contexts, social support can reduce the levels of emotional exhaustion, depersonalization, and reduced performance that are often associated with burnout. Through positive interactions with family, friends, or coworkers, individuals can feel supported and appreciated, creating an environment that supports psychological well-being. Social support can also play a role in increasing feelings of self-control and self-efficacy, reducing feelings of isolation, and providing constructive solutions to work problems. However, it is important to remember that the quality of social support and individual perceptions of that support may vary. In some cases, inadequate social support or an individual’s questionable perception of support can reduce its effectiveness. Therefore, planning and implementing appropriate social support strategies needs to take individual needs and preferences into account.

## Discussion

Social support, which includes emotional help, instrumental support, and social engagement, can have a significant impact on a person’s level of burnout. Burnout, which is a state of physical, emotional and mental exhaustion that can arise due to chronic stress, especially related to work or life demands, can be minimized and overcome through the important role of social support. Social support can play a role in reducing the risk and impact of burnout by providing a channel for sharing burdens, improving psychological well-being, and facilitating more effective coping strategies. Through the exchange of experiences and resources, social support can form networks of trust and solidarity, create an environment that supports personal and professional growth, and provide protection against stress that can lead to burnout. Therefore, understanding and utilizing the positive role of social support can be a key step in preventing and treating burnout holistically. and the impact of burnout in several ways:

### Emotional Support (Emotional Support)

Emotional support from friends, family, or coworkers can help individuals feel heard and understood. Knowing that there are people who care and are willing to listen can reduce feelings of isolation that are often associated with burnout.

### Instrumental Support

Support in the form of practical help, such as help with household chores or work assignments, can reduce the burden that may trigger burnout.

### Informational Support

Providing relevant information or advice can help individuals overcome problems that may be a source of stress and burnout.

### Perception of Social Support

The importance of not only getting social support but also feeling supported. Individuals’ perceptions of the level of social support they receive can influence their well-being.

### Stress Reduction

Social support can act as a buffer against the effects of stress, helping individuals handle the pressure better.

However, it is important to remember that social support cannot always prevent burnout completely. Several other factors, such as excessive work demands, lack of control, and lack of organizational support, can also play a role in the emergence of burnout. Therefore, the best approach may involve a combination of social support with organizational changes that support employee well-being.

## Conclusions And Suggestions

Social support can be considered a form of “natural protection” against burnout, and efforts to strengthen social support networks can be an effective strategy in preventing and treating burnout at various levels of life and professions.

## Data Availability

All data produced are available online at nursingcecentral

## Reviewer’s Advice

The author entrusts the complete review of our article to the manager, and any necessary corrections based on feedback from the review team are communicated back to us.

